# Long-Term SARS-CoV-2-Specific Immune and Inflammatory Responses Across a Clinically Diverse Cohort of Individuals Recovering from COVID-19

**DOI:** 10.1101/2021.02.26.21252308

**Authors:** Michael J. Peluso, Amelia N. Deitchman, Leonel Torres, Nikita S. Iyer, Christopher C. Nixon, Sadie E. Munter, Joanna Donatelli, Cassandra Thanh, Saki Takahashi, Jill Hakim, Keirstinne Turcios, Owen Janson, Rebecca Hoh, Viva Tai, Yanel Hernandez, Emily Fehrman, Matthew A. Spinelli, Monica Gandhi, Lan Trinh, Terri Wrin, Christos J. Petropoulos, Francesca T. Aweeka, Isabel Rodriguez-Barraquer, J. Daniel Kelly, Jeffrey N. Martin, Steven G. Deeks, Bryan Greenhouse, Rachel L. Rutishauser, Timothy J. Henrich

## Abstract

A detailed understanding of long-term SARS-CoV-2-specific T cell responses and their relationship to humoral immunity and markers of inflammation in diverse groups of individuals representing the spectrum of COVID-19 illness and recovery is urgently needed. Data are also lacking as to whether and how adaptive immune and inflammatory responses differ in individuals that experience persistent symptomatic sequelae months following acute infection compared to those with complete, rapid recovery. We measured SARS-CoV-2-specific T cell responses, soluble markers of inflammation, and antibody levels and neutralization capacity longitudinally up to 9 months following infection in a diverse group of 70 individuals with PCR-confirmed SARS-CoV-2 infection. The participants had varying degrees of initial disease severity and were enrolled in the northern California Long-term Impact of Infection with Novel Coronavirus (LIINC) cohort. Adaptive T cell responses remained remarkably stable in all participants across disease severity during the entire study interval. Whereas the magnitude of the early CD4+ T cell immune response is determined by the severity of initial infection (participants requiring hospitalization or intensive care), pre-existing lung disease was significantly associated with higher long-term SARS-CoV2-specific CD8+ T cell responses, independent of initial disease severity or age. Neutralizing antibody levels were strongly correlated with SARS-CoV-2-specific CD4+ T but not CD8+ T cell responses. Importantly, we did not identify substantial differences in long-term virus-specific T cell or antibody responses between participants with and without COVID-19-related symptoms that persist months after initial infection.

## INTRODUCTION

There is an ongoing and urgent need to understand the development and persistence of natural immunity in individuals who have recovered from SARS-CoV-2 infection. Until vaccination becomes widely available and accessible, many previously infected persons will continue to rely upon natural immunity to protect against re-infection. A detailed understanding of T cell responses to natural infection could also inform our expectations for the magnitude and durability of these responses after vaccination.

Most people generate detectable SARS-CoV-2-specific CD4+ and CD8+ T cell responses following natural infection with the virus (Grifoni et al. 2020; Peng et al. 2020; Sekine et al. 2020; Rydyznski Moderbacher et al. 2020; Braun et al. 2020; Zhou et al., n.d.; Breton et al. 2021; Dan et al. 2021). However, current understanding of the factors associated with the magnitude and long-term persistence of the cellular immune response and its relationship to clinical outcomes, humoral responses, and soluble markers of inflammation remain limited. In particular, there is a need to understand these responses in diverse groups of individuals representing the spectrum of COVID-19 illness and recovery. To date, many cohort studies describing SARS-CoV-2 memory T cell responses have selectively included mostly women (Peng et al. 2020), younger individuals with few comorbidities and mild disease (Grifoni et al. 2020), and white study participants (Breton et al. 2021), therefore limiting data on the immunologic characteristics of specific groups that have been disproportionately affected by COVID-19 (Breton et al. 2021; Richardson et al. 2020; Chamie et al. 2020; Figueroa et al. 2020; Wadhera et al. 2020). Furthermore, there is growing interest in understanding whether important immunologic differences may exist among groups experiencing rapid versus prolonged COVID-19 recovery, but data from this latter group are lacking (Hellmuth et al. 2021; Tenforde et al. 2020; Carfì et al. 2020; Drew et al. 2020; Huang et al. 2021; Datta, Talwar, and Lee 2020).

To address these important issues, we measured SARS-CoV-2-specific T cell responses, soluble markers of inflammation, or antibody levels and neutralization capacity longitudinally up to 8.9 months following infection in a diverse group of 70 individuals with PCR-confirmed SARS-CoV-2 infection with varying degrees of initial disease severity in northern California enrolled in the Long-term Impact of Infection with Novel Coronavirus (LIINC) cohort. We demonstrate that, while the magnitude of the initial cellular immune response is determined by the severity of initial infection, these responses remain remarkably stable for up to 8 months. Whereas the magnitude of the early CD4+ T cell immune response is determined by the severity of initial infection (participants requiring hospitalization or intensive care), pre-existing lung disease was significantly associated with higher long-term SARS-CoV2-specific CD8 T cell responses, independent of initial disease severity or age. Neutralizing antibody levels were strongly correlated with SARS-CoV-2-specific CD4+ T but not CD8+ T cell responses. Importantly, we did not identify substantial differences in long-term virus-specific T cell or antibody responses between participants with and without COVID-19-related symptoms that persist months after initial infection.

## RESULTS

### Characterization of a clinically diverse COVID-19 cohort over eight months of recovery

In order to evaluate adaptive immune and inflammatory responses over a range of COVID-19 presentations, we selected 70 cohort participants that represented a wide range of initial disease presentations, from those with no or mild symptoms to those requiring hospitalization or treatment in an intensive care unit (ICU). One participant had antibody and inflammatory marker data but insufficient cell viability for T cell analyses. We prioritized inclusion of participants enrolled during early recovery (within 40 days following onset of symptoms) and those with samples available at later time points after symptom onset. Peripheral blood mononuclear cells (PMBCs), plasma and serum were collected longitudinally between one and 8 months after symptom onset.

Overall, 48.6% of participants were female, 25.7% identified as Latino or Latina (Latinx) and 39 55.7% identified as white (non-Latinx) as shown in **Table 1**. 25.7% participants were hospitalized (all but two required supplemental oxygen), and 14.3% reported receiving ICU care. A significantly higher proportion of males than females were hospitalized (38.9% vs 11.8%; P=0.02) and all participants who received ICU care were male (P<0.01). A significantly higher proportion of hospitalized participants (61.1%) identified as Latinx (P<0.001). The majority of participants did not have underlying medical comorbidities, but 18.6% were previously diagnosed with lung disease (*e.g*., asthma, chronic obstructive pulmonary disease, etc.), 12.9% with hypertension and 8.6% with diabetes mellitus (DM). Given the large number of individuals enrolled prior to widespread availability of COVID-19-specific therapies, only 4.3% reported having received remdesivir, 11.4% received hydroxychloroquine with or without azithromycin, and one participant each reported receiving systemic corticosteroids and convalescent plasma.

**Table 1.** Participant Demographics, Co-Morbidities and Clinical Presentations.

|  | All<br>Participants | Sex |  | Hospitalized |  |
| --- | --- | --- | --- | --- | --- |
|  |  | Female | Male | Yes | No |
| N | 70<sup>a</sup> | 34 | 36 | 18 | 52 |
| Age (Median, IQR) | 43 (36, 53) | 43 (36.8, 53) | 44.5 (36, 55.3) | 50.5 (40.3, 56.8) | 40 (36, 53) |
| Female (N, %<sup>b</sup>) | 34 (48.6) | - | - | **4 (22.2)\*** | **30 (57.7)** |
| Race/Ethnicity (N, %) |  |  |  |  |  |
| Latinx | 18 (25.7) | 6 (17.6) | 12 (33.3) | **11 (61.1)\*\*\*** | **5 (17.9)** |
| White (Non Latinx) | 39 (55.7) | 23 (67.6) | 16 (44.4) | 3 (16.7) | 19 (67.9) |
| Black | 3 (4.3) | 3 (8.8) | 0 (0) | 1 (5.6) | 2 (7.1) |
| Asian | 10 (14.3) | 2 (5.9) | 8 (22.2) | 3 (16.7) | 2 (7.1) |
| Days from Symptom Onset to 1<sup>st</sup> Sample (Median, IQR) | 53 (38, 64.5) | 50 (37.8, 63.3) | 55 (41.3, 70) | 73 (51.8, 89.8) | 48 (37.3, 60.8) |
| ICU Admission (N, %) | 10 (14.3) | **0 (0)\*\*** | **10 (100)** | - | - |
| Underlying Medical Condition (N, %) |  |  |  |  |  |
| Lung Disease | 13 (18.6) | 9 (26.5) | 4 (11.1) | 5 (27.8) | 8 (15.4) |
| Autoimmune Disease | 3 (4.3) | 3 (8.8) | 0 (0) | 0 (0) | 3 (5.8) |
| Hypertension | 9 (12.9) | 6 (17.6) | 3 (8.3) | 4 (22.2) | 5 (9.6) |
| Cancer | 1 (1.4) | 1 (2.9) | 0 (0) | 0 (0) | 1 (1.9) |
| Diabetes Mellitus | 6 (8.6) | 2 (5.9) | 4 (11.1) | **5 (27.8)\*\*** | **1 (1.9)** |
| Symptoms During Acute Infection (N, %) | 68 (97.1) | 32 (94.1) | 36 (100) | 18 (100) | 50 (96.2) |
| Fever/Chills | 56 (80) | 24 (70.6) | 32 (88.9) | 17 (94.4) | 39 (75) |
| Cough/SOB | 61 (87.1) | 28 (82.4) | 33 (91.7) | 17 (94.4) | 44 (84.6) |
| Sore Throat/Runny Nose | 46 (65.7) | 22 (64.7) | 24 (66.7) | 10 (55.6) | 36 (69.2) |
| Cardiac (chest pain, palpitations) | 7 (10) | 2 (5.9) | 5 (13.9) | 3 (16.7) | 4 (1.9) |
| Neurological/Cognitive | 49 (70) | 26 (76.5) | 23 (63.9) | 13 (72.2) | 36 (69.2) |
| Fatigue | 63 (90) | 31 (91.2) | 32 (88.9) | 16 (88.9) | 47 (90.4) |
| Smell/Taste changes | 52 (74.3) | 28 (82.4) | 24 (66.7) | 13 (72.2) | 39 (75) |
| Gastrointestinal symptoms | 43 (61.4) | 24 (70.6) | 19 (52.8) | **14 (77.8)\*** | **29 (55.8)** |
| Muscle aches | 48 (68.6) | 34 (70.6) | 24 (66.7) | 14 (77.8) | 34 (65.4) |
| Persistent Symptoms at time of first sample (N, %) | (45.8) | 18 (52.9) | 14 (38.9) | 9 (50) | 23 (44.2) |
| Fever/Chills | 0 (0) | 0 (0) | 0 (0) | 0 (0) | 0 (0) |
| Cough/SOB | 15 (21.4) | 8 (23.6) | 7 (19.4) | 6 (33.3) | 9 (17.3) |
| Sore Throat/Runny Nose | 3 (4.3) | 0 (0) | 3 (8.3) | 1 (5.6) | 2 (3.8) |
| Cardiac (chest pain, palpitations) | 2 (2.9) | 1 (2.94) | 1 (2.8) | 1 (5.6) | 1 (1.9) |
| Neurological | 14 (20) | 6 (17.6) | 8 (22.2) | 5 (27.8) | 9 (17.3) |
| Fatigue | 11 (15.7) | 7 (20.6) | 4 (11.1) | 4 (22.2) | 7 (13.5) |
| Smell/Taste changes | 15 (21.4) | 9 (26.5) | 6 (16.7) | 6 (33.3) | 9 (17.3) |
| Gastrointestinal symptoms | 7 (10) | 5 (14.7) | 2 (5.6) | 3 (16.7) | 4 (7.7) |
| Muscle aches | 6 (8.6) | 4 (11.8) | 2 (5.6) | 3 (16.7) | 3 (5.8) |
| Days from Symptom Onset to 4<sup>th</sup> Sample (Median, IQR) | 125 (120, 133) | 127 (122, 132) | 123 (116, 134) | 125.5 (120.3, 134.5) | 125 (120, 125) |
| Persistent Symptoms at 4<sup>th</sup> Sample (N, %) | 24 (36.4) | 15 (45.5) | 9 (27.3) | 8 (50) | 11 (40.7) |

(\*) P<0.05, (\*\*) P<0.01, (\*\*\*) P<0.001 by Fisher's Exact Test for within column comparisons; SOB = shortness of breath
<sup>a</sup> One participant had antibody and inflammatory marker data but insufficient cell viability for T cell analyses
<sup>b</sup> Percentage of participants for the column N are shown throughout the table

Ninety seven percent of participants had COVID-19-related symptoms at the time of or immediately following their initial diagnosis, and 45.8% had persistence of at least one COVID-19-attributed symptom during the initial convalescent visit which was a median of 53 days since symptom onset (or initial time of PCR positivity for asymptomatic individuals; **Table 1**). 36.4% had COVID-19-related symptoms approximately 4 months after initial illness (median 125 days since initial symptom onset). Although not statistically significant, a higher proportion of females than males reported persistent symptoms at the initial and four-month study visits (52.9% versus 38.9%, and 45.5% versus 27.3%, respectively), despite lower hospitalization rates and similar time to study enrollment. Pulmonary symptoms including cough and shortness of breath, fever and loss or changes in smell or taste, fatigue and neurological symptoms (including headache, difficulties with concentration, attention, brain fog, neuropathies) were the most commonly experienced symptoms during acute infection (all reported in >70% of participants) and during the first study visit (**Table 1**).

### SARS-CoV-2-specific T cell responses are stable over 8 months following initial infection

We applied two different methods to ascertain the frequency of SARS-CoV-2-specific CD4+ and CD8+ T cells in the peripheral blood after stimulation with Spike- or Nucleocapsid-derived peptide pools: the activation induced marker (AIM) assay, which measures co-expression of OX40 and CD137 (4-1BB) on CD4+ T cells or co-expression of CD69 and CD137 on CD8+ T cells (Grifoni et al. 2020) (gating strategy shown in **Supplementary Figure 1**), and an intracellular cytokine staining (ICS) assay to ascertain the frequencies of CD8+ and CD4+ T cells expressing interferon (IFN)γ, co-expressing tumor necrosis factor (TNF)α and IFNγ, and IFNγ-cells expressing the degranulation marker CD107a. The percentage of CD8+ T cells identified in the ICS assay by the expression of CD107a and/or IFNγ that co-express Granzyme B was also measured (gating strategy shown in **Supplementary Figure 2**). Overall, 192 unique sample time points across 69 participants ranging from 26 to 266 days (8.9 months) after onset of symptoms were tested and yielded interpretable results from either AIM or ICS assays. A large majority of participants had detectable Spike (S) or Nucleocapsid (N) protein SARS-CoV-2-specific non-naïve (memory) CD4+ (100%) and CD8+ (95.7%) T cell responses greater than the upper interquartile range of five historical blood banked samples collected prior to November 2019 at at least one time point over the entire study period by either AIM or ICS assays (**Supplementary Table 1**). An upper quartile cut-off was used given potential for noise in certain pre-COVID samples which may have been due prior coronavirus infection cross-reactivity as has previously been shown (Sekine et al. 2020). A median of 23.1 (IQR 10.3, 36.6)% of IFNγ+ CD8+ T cells (N and S-specific) and 61.5 (IQR 44.4, 76.0)% of IFNγ+ CD4+ T cells (N- and S-specific) produced TNFα across all timepoints sampled (P<0.001).

The frequency of SARS-CoV-2 S- and N-specific memory CD4+ and CD8+ T cells (measured as AIM+, or ICS+: IFNγ+, IFNγ+/TNFα+, or IFNγ-/CD107a+) for the most part did not significantly change over the sampling period (*i.e.,* slopes were not different from zero in linear mixed effects models). The exception to this was a modest decrease in the frequency of CD4+ T cells expressing IFNγ+ in response to N peptide stimulation (−0.0065 log_2_ change in percentage per day, P=0.011). These data demonstrate long-term stability of SARS-CoV-2-specific T cell responses in our cohort (**Figure 1; Supplementary Figure 3**).

**Figure 1.**
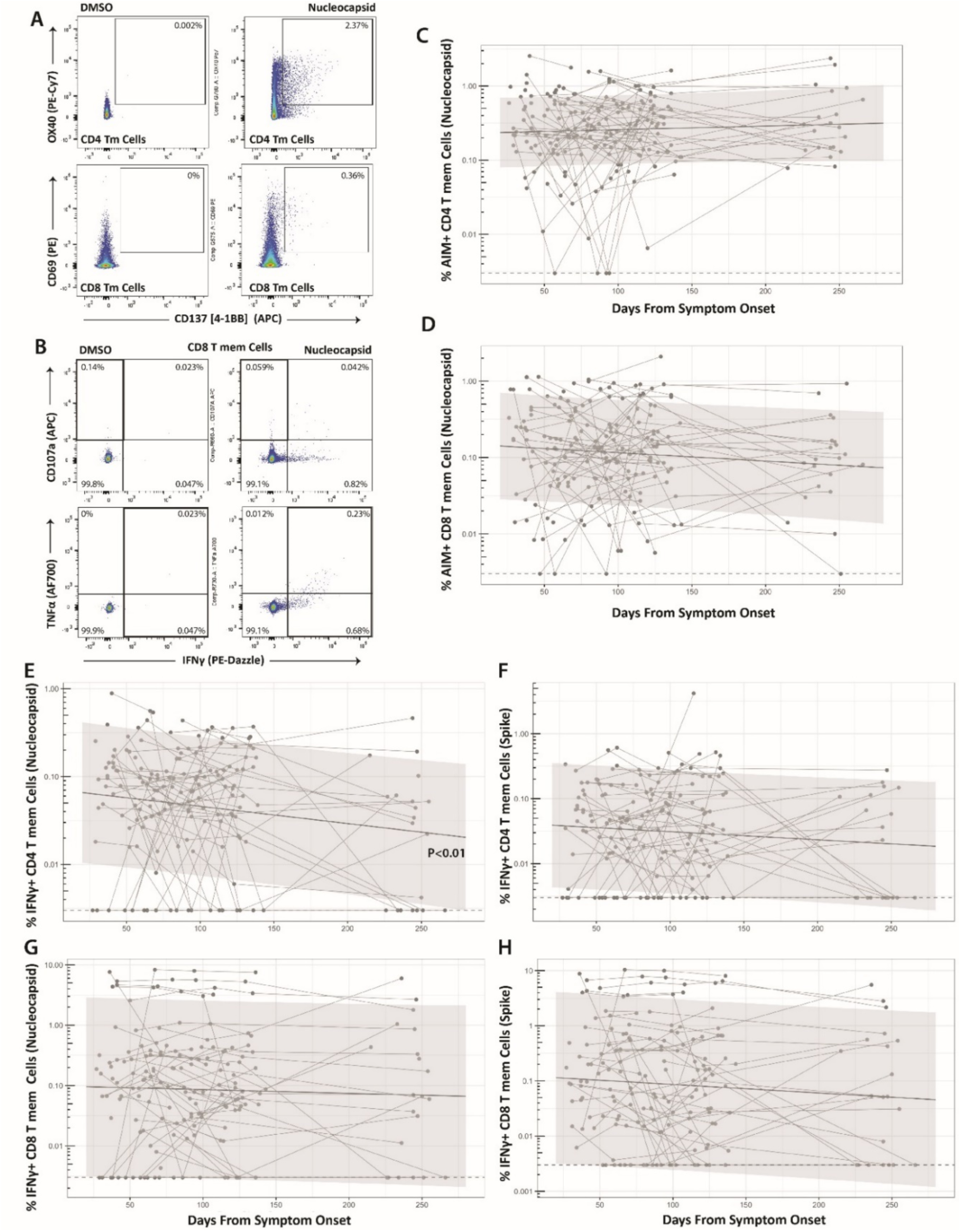
Long-term durability of SARS-CoV-2-specific CD4 and CD8 cell responses. Gating strategy for identifying SARS-CoV-2-specific memory T cell responses in the activation-induced marker (AIM) (**A**) and intracellular cytokine staining (ICS) (**B**) assays. Percentage of AIM+ N-specific CD4+ (**C**) and CD8+ (**D**) T cell responses over time (S-specific responses were similar and are shown in Supplementary Figure 3). Percentage of IFNγ+ Nucleocapsid (N)- or Spike (S)-specific CD4 (**E-F**) and CD8 (**G-H**) T cell responses over time. Points and connecting lines represent raw data for each individual. Solid line and shaded region represent the median model prediction and 95% prediction interval from linear mixed effects modeling including individual effects. Dashed lines represent assay limits of detection. T mem = T memory cells.

### Differences in SARS-CoV-2-specific T cell responses across demographic and disease factors

We next asked whether the frequency of SARS-CoV-2-specific CD4+ and CD8+ T cells (AIM+, IFNγ+, IFNγ+/TNFα+, and IFNγ-/cd107a) were impacted by various clinical and demographic factors. We first employed cross-sectional analyses at two unique study time points, one early and one late dring study collections (median 53 days after symptom onset [IQR 38-64.5] versus median 123 days [IQR 115-130.5], respectively). We then performed longitudinal analyses using linear mixed effects modeling over all time points, with days from onset of symptoms and individual factors (*i.e.* sex, race/ethnicity, age, COVID-19-related hospitalization, prior ICU care, and the presence of persistent COVID-19-related symptoms) as model parameters. Longitudinal models incorporated random variability for each participant.

We observed differences in the frequency of SARS-CoV-2-specific memory CD4+ and/or CD8+ T cell responses according to several demographic and clinical parameters in both cross-sectional (early and late time points) and longitudinal linear mixed effects models. We highlight key differences in IFNγ-expressing cell populations identified by the ICS assay in **Figure 2** (differences in polyfunctional responses as determined by co-expression of IFNγ and TNFα followed a similar pattern and are shown in **Supplementary Figure 4**). Participants hospitalized during acute infection had significantly higher frequencies of N and S-specific IFNγ+ CD4+ T cells at the early analysis time point and S-specific IFNγ+ CD4+ T cells at the late analysis time point compared to non-hospitalized participants (**Figure 2A**). We also observed higher N and S IFNγ+ CD4+ responses in hospitalized participants mixed effects models across all longitudinal data points through the last collection time point at study month 8 (P= 0.025 and 0.068; **Figure 2B**). The subset of hospitalized participants who were admitted to the ICU had significantly higher frequencies of N-specific IFNγ+ CD4+ T cells at both cross-sectional early and late analysis time points compared to participants that did not require ICU care, regardless of hospitalization (**Figure 2C**). N-specific IFNγ+ CD4+ T cell responses were also significantly higher for participants receiving ICU care across all time points in linear effects modeling (P=0.024).

**Figure 2.**
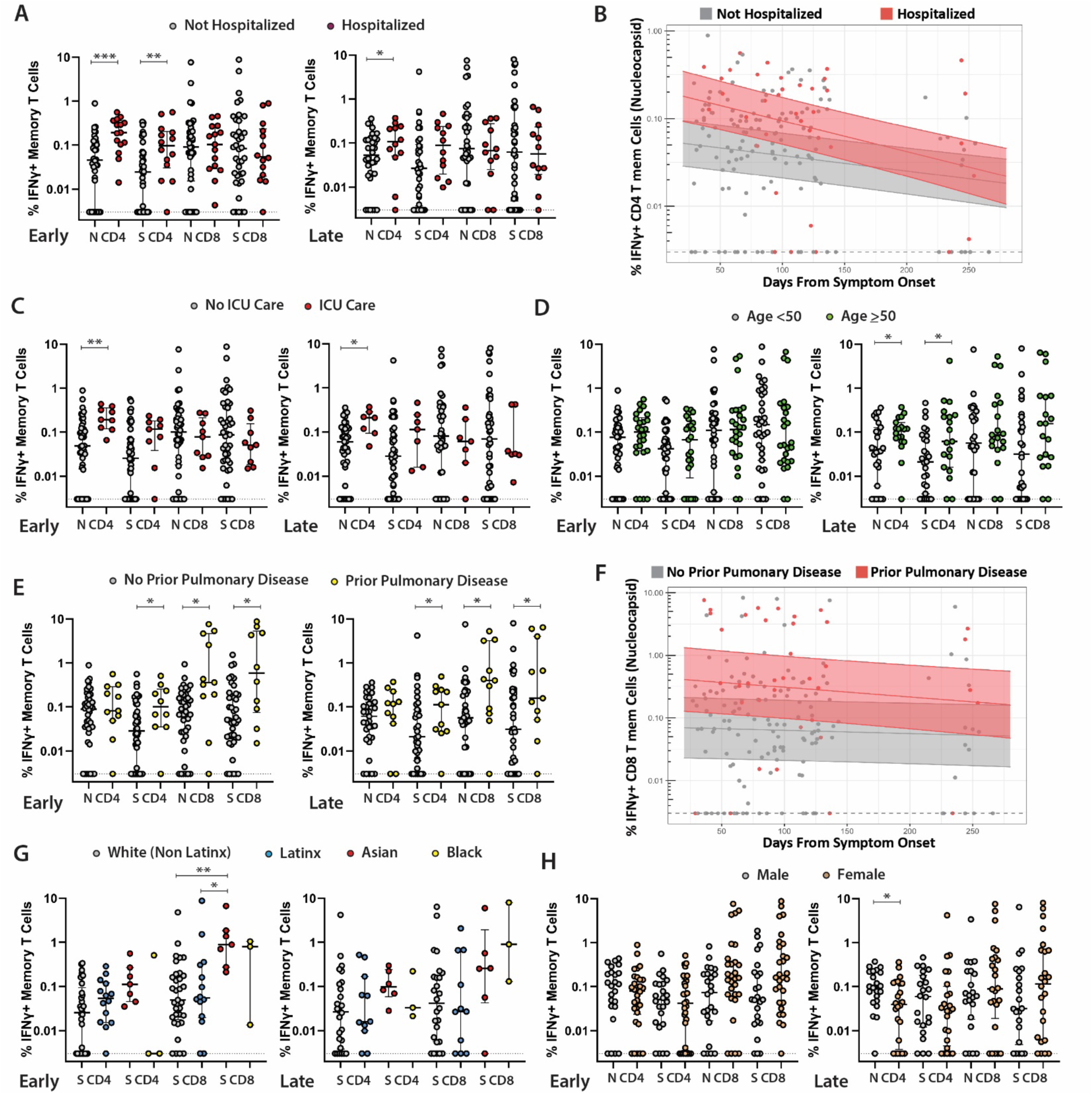
Relationships between SARS-CoV-2-specific T cell responses and participant demographic and clinical factors. Frequency of SARS-CoV-2-specific CD4+ T cells as measured by the ICS assay in study participants who were (**A**) hospitalized versus not hospitalized at early (left, median 53 days after onset of symptoms) versus late (right, median 123 days from onset of symptoms) cross-sectional analysis timepoints. (**B**) Longitudinal frequency of IFNγ+ memory CD4+ T cells in hospitalized versus non-hospitalized participants. (**C**) Frequency of SARS-CoV-2-specific CD4 T cells as measured by the ICS assay in study participants who required ICU care. (**D**) Frequency of SARS-CoV-2-specific CD4+ T cells as measured by the ICS assay in study participants <50 versus ≥50 years of age at early and late cross-sectional analysis timepoints. Cross-sectional (**E**) and longitudinal (**F**) frequency of SARS-CoV-2-specific CD8 T cells in study participants who had diagnosed pulmonary disease prior to infection. (**G**) Frequency of SARS-CoV-2-specific CD8 T cells amongst participants identifying as white (non-Latinx), Latinx, Asian or Black. All data points are shown as individual points. For **A**, **C**, **D**, **E**, **G**, **H**: Bars and lines in cross sectional data represent median values and interquartile ranges; (*) P<0.05, (**) P<0.01, (***) P<0.001 by non-parametric Kruskal-Wallis tests with Dunn adjustment for analyses incorporating comparisons across more than two variables. For **B**, **F**: solid line and shaded region represent the median model prediction (including individual effects) and 50% prediction interval. T mem = T memory cells.

While we did not observe differences between participants <50 or ≥50 years of age at the early analysis time point, those aged ≥50 years had significantly higher percentages of N and S-specific IFNγ+ CD4+ T cells at the late analysis time point (**Figure 2D; Supplementary Figure 4**). Interestingly, participants with a pre-existing history of pulmonary disease had significantly higher percentages of S-specific IFNγ+ CD4+ T cells and S-specific IFNγ+ CD8+ T cells at both early and late analysis time points (**Figure 2E**; **Supplementary Figure 4**). We observed significantly higher N and S IFNγ+ CD8+ (P=0.018, 0.016; **Figure 2F**) and S IFNγ+ CD4+ (P=0.024) T cell responses in participants with prior pulmonary disease in mixed effects models across all longitudinal data points through the last collection time point at study month 8.

A significantly higher frequency of S-specific IFNγ+ CD8+ T cells was also observed in Asian compared with white (non-Latinx) participants (**Figure 2G**) at the first analysis time point only, but there were no differences between Latinx and white (non-Latinx) participants at either the early or late cross-sectional analysis time points by the ICS assay. In contrast, we observed a significantly higher percentage of S AIM+ CD4+ T cells in Latinx versus white (not-Latinx) participants at the early analysis time point (P=0.025), but not at the later time point (P=0.97). Linear mixed effects modeling also revealed a higher overall percentage of N AIM+ CD4+ T cells (P=0.022) and S AIM+ CD8+ T cells (P=0.003) in Latinx versus white (non-Latinx) participants across all longitudinal data points. Despite higher overall levels, the percentage of S AIM+ CD8+ T cells declined in the Latinx but not in the white non-Latinx population (P=0.006; **Supplementary Figure 5**).

We did not find significant differences in the frequency of SARS-CoV-2-specific CD4+ and CD8+ T cells by ICS assay between males and females or between individuals who did or did not have persistent symptoms at the early or late cross-sectional time points or from linear mixed effects models across all time points (all P>0.05); **Figure 2H**). Importantly, we did not observe any major differences between T cell immune responses in those with or without persistent symptoms at either the initial study time point or approximately 4 months following onset of acute infection in cross-sectional or longitudinal analyses (**Supplementary Figure 6**). We observed no differences between any demographic or clinical comparator groups at either the first or last cross-sectional analysis time points for S- or N-specific IFNγ-/CD107a+ CD4+ or CD8+ T cell responses. Overall, a majority of SARS-CoV2-specific CD8+ T cells that expressed either IFNγ or CD0107a at the early and late memory cross-sectional analysis time points expressed

Granzyme B (median 66.7% and 77.3% for N- and S-responsive cells at the early analysis time point and 55.6% and 67.1% at the late analysis time point within both IFNγ+ and IFNγ-CD107a+ cells). No significant differences between early and late time points within N- and S-specific T cells were observed (all P>0.68).

### Interactions and independent associations between T cell responses and demographic and clinical factors

In order to determine the relationships between significant covariates identified in univariate cross-sectional analyses as above (COVID-19-related hospitalization, pre-existing lung disease and age), we performed linear regression modeling using Log_2_ transformed data for T cell responses at the initial and last analysis time points and performed tests of interactions between covariates. Differences in N and S-specific IFNγ+ CD4+ T cell responses at the first time point and S-specific IFNγ+ CD4+ T cell response at the last time point between hospitalized and non hospitalized participants remained significant at the first time point when adjusted for age (all P<0.042). Furthermore, increased percentages of N and S-specific IFNγ+ CD8+ T cells at the first time point and N-specific IFNγ+ CD8+ T cells at the last time point observed in participants with pre-existing lung disease were significant in regression models adjusted for age (all P<0.042). Similarly, all increases in N and S-specific IFNγ+ CD8+ T cell responses observed in those with pre-existing lung disease in univariate analyses remained significant when adjusted for prior hospitalization (all P<0.024).

We identified significant interactions between hospitalization and age for S-specific IFNγ+ CD4+ T cells at the last analysis time point (P=0.007), demonstrating that with more advanced age, there were lower percentages of IFNγ-producing CD4+ T cells in hospitalized participants even though the percentage of IFNγ+ CD4+ T cells were generally higher for age ≥50 and hospitalization in univariate analyses described above. Interestingly, participants with a prior history of lung disease and COVID-19 related hospitalization had lower N-specific IFNγ+ CD8+ T cells at the last analysis time point (P=0.024) despite higher levels observed in those with a pre-existing lung disease only.

### SARS-CoV-2 specific antibody responses and neutralization capacity strongly correlate with CD4+ T cell responses

We quantified IgG antibody titers to full-length SARS-CoV-2 N, S, and N fragment (N.361), and the S receptor binding domain (RBD) proteins using an in-house Luminex assay. We also tested neutralizing antibody (NAb) responses using pseudoviruses expressing the Spike protein in the presence of autologous sera from approximately 4 months post-symptom onset (median 125 days [IQR 120-133]) across the entire study population (N = 66; 4 participants did not have NAb data available). Consistent with data that we and others (Seow et al. 2020; Long et al. 2020; Gudbjartsson et al. 2020; Naaber et al. 2020; Y. Chen et al. 2020; X. Chen et al. 2020; Kowitdamrong et al. 2020; Lei et al. 2020; Zhao et al. 2020; Gaebler et al. 2021) have previously demonstrated, N, S and RBD-specific antibody levels remained stable over time (**Supplemental Figure 7**). Higher antibody titers were observed at the early and late memory timepoints in those who were hospitalized (including the subgroup requiring ICU level care; data not shown) and those who identified as Latinx, and at the late memory timepoint only amongst participants ≥ 50 years of age (N-specific responses only for the latter) (all P <0.035; **Figure 3A-C**). In linear regression modeling of antibody responses to determine the independent relationships between covariates of interest, we observed that increased N and RBD antibody levels observed in univariate analyses at the first time point in hospitalized participants remained statistically significant when adjusted for Latinx race/ethnicity (P=0.02, 0.05). However, the increases in antibody levels observed in Latinx participants were not significant in adjusted analyses, suggesting that differences in antibody responses are primarily driven by initial disease severity, rather than by demographic factors.

**Figure 3.**
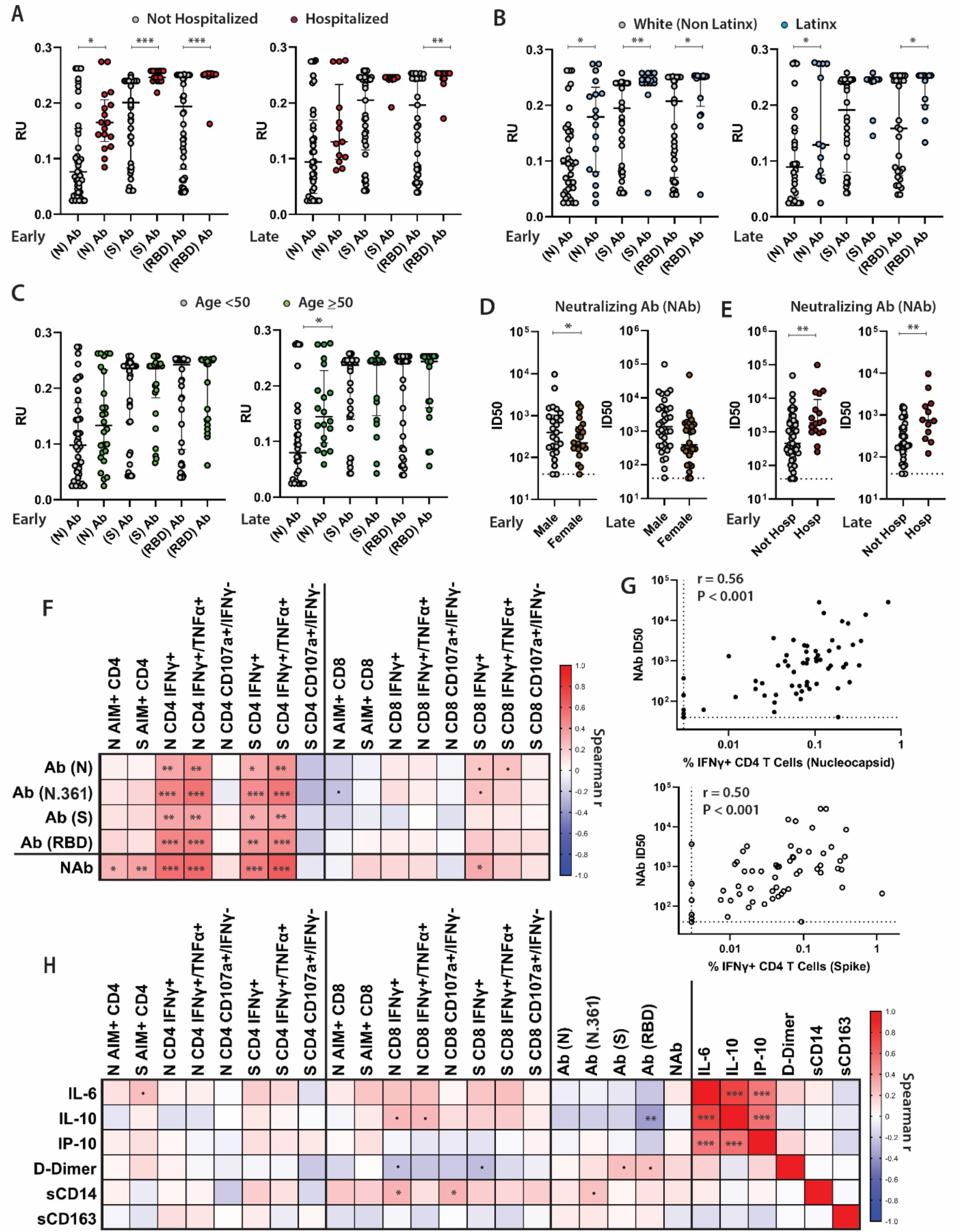
SARS-CoV-2-specific antibody levels and neutralization responses, soluble markers of inflammation, and relationships with T cell responses. Differences in the levels of nucleocapsid (N), spike (S), and receptor binding domain (RBD)-specific antibody responses for participants grouped by age (**A**), hospitalization during acute infection (**B**), and Latinx versus white-non Latinx race/ethnicity (**C**) at the early and late memory analysis time points. Infectious dose, 50% (ID50) values from neutralization assays in cross sectional neutralization antibody (NAb) analyses of participants grouped by sex (**D**) and hospitalization during acute infection (**E**). Spearman correlation matrix heatmap (r values) between weighted average antibody and T cell response across all time points (**F**) and individual correlation plots between NAb ID50 and the percentage of CD4 T cells co-expressing IFNγ+ in response to N and S peptide pools (**G**). (**H**) Spearman correlation matrix heatmap incorporating soluble markers of inflammation and T cell and antibody responses. All data points are shown on dot plots. Dashed lines represent assay limits of detection. Solid bars and lines represent median values and interquartile ranges. (.) P<0.1, (*) P<0.05, (**) P<0.01, (***) P<0.001 by non-parametric analyses.

Neutralizing antibody responses were modestly lower in female participants at the first study time point (P=0.013), but there were no significant differences observed at the last cross-sectional analysis time point (**Figure 3D**). In longitudinal analyses, N, N fragment (N.361), S and RBD-specific antibody levels were higher across all time points for hospitalized participants (P=0.024, 0.012, 0.014, 0.043, respectively).

In order to explore relationships between T cell responses and antibody levels and soluble inflammatory markers, we generated Spearman correlation matrices (**Figure 3F**). Given the variation in time from initial symptoms to sample collection times and the number of sample time points for each participant, correlation analyses were performed using weighted averages for all T cell, antibody and cytokine data across all time-points. N, N.361, S and RBD-specific antibody levels were significantly correlated with N- and S-specific CD4+ T cell responses (as measured by expression IFNγ as well as dual expression of IFNγ and TNFα), but much less so with CD8+ T cell responses (**Figure 3F** and **Supplementary Figure 8**). Neutralizing antibody responses were most strongly correlated with S- and N-specific CD4+ T cell IFNγ+ and IFNγ+/TNFα+ ICS results, and also correlated with S- and N-specific AIM responses as shown in **Figure 3F, G**.

### Relationships between soluble markers of inflammation and immune responses

We measured longitudinal circulating levels of IL-6, IL-10, IP-10, D-Dimer, sCD14 and sCD163 in a random subset of 57 individuals through study month 4 (**Supplementary Figure 9**). There were no significant changes in the levels of soluble markers over time, with the exception of a modest decline in sCD14 (P=0.006). No significant differences were observed between demographic and clinical factor groups in cross sectional analysis at the first and last analysis time points. Interestingly, in longitudinal analyses we observed modestly lower IL-6 and IL-10 levels in hospitalized participants (P=0.007 and P=0.047, respectively) and those who received ICU care (P=0.024 and P=0.043, respectively) across all time points. IP-10 levels were modestly higher in the hospitalized across timepoints (P=0.028) and in those with persistent symptoms four months following acute illness (P=0.034).

Spearman correlation analysis was also performed for the soluble markers of inflammation and T cell and antibody responses using weighted averages as in **Figure 3H**. Overall, there were modest positive associations between IL-1, IL-10 and sCD14 levels with the frequency of CD8+ T cell responses as measured by ICS and negative association with D-Dimer levels with CD8+ T cell ICS responses, but many of these associations did not reach statistical significance (**Figure 3H**, **Supplementary Figure 7**). We did observe a negative correlation between IL-10 levels and RBD-specific antibody responses, however. IL-6, IL-10, and IP-10 were strongly positively correlated with each other but not with D-Dimer, sCD14 or sCD163.

## DISCUSSION

We demonstrate in a well-characterized, diverse cohort with long-term follow-up that adaptive immune responses are relatively stable over 8 months following infection with SARS-CoV-2. While our data do not inform on the clinical protection of SARS-CoV-2-specific immunity, the stability of these responses over time is certainly promising for the durability of natural and potentially vaccine-induced immunity. These findings greatly expand upon descriptions of T cell immunity in cohorts with more homogeneity in participant demographics and/or disease severity (Braun et al. 2020; Peng et al. 2020; Rydyznski Moderbacher et al. 2020; Grifoni et al. 2020; Breton et al. 2021; Dan et al. 2021). For example, we observed potentially important differences in CD4+ and CD8+ T cell adaptive immune responses across various groups and differences during both early and late recovery (median of 53 days and 123 after onset of symptoms).

More specifically, those requiring hospitalization or intensive care during acute infection had higher levels of memory CD4+ T cell responses at the earlier convalescent time point. Whereas the higher percentage of SARS-CoV-2-specific CD4+ T cell responses initially observed in hospitalized participants converged with those in participants who were not hospitalized approximately 4 months following acute infection similar to one prior report (Dan et al. 2021), CD4+ T cell responses remained elevated in those who had required ICU level care. CD4+ T cell responses generally mirrored nucleocapsid, spike and receptor binding domain antibody responses. In contrast, CD8+ T cell responses were less strongly correlated with antibody responses, but we identified persistently higher virus-specific CD8+ T cell responses in the subset of participants with pre-existing pulmonary disease. These differences remained significant after adjusting for prior hospitalization and age. In addition, we observed an interesting interaction in that those with pre-existing lung disease had *lower* SARS-CoV-2-specific CD8+ T cell responses if hospitalized. As a result, our results suggest that the relationships between various demographic and clinical factors are complex and will require large, diverse, and well-curated cohorts to more fully understand relationships between long-term immunity and different components of the immune response.

Older age is a major factor associated with severe COVID-19 and death and there is ongoing concern that older individuals will experience more rapid decline in viral-specific immunity. Interestingly, we observed minimal age-related differences in the SARS-CoV-2-specific CD4+ T cell response in early convalescence, and a significantly higher response in older individuals months later. There were no significant age-related differences in CD8+ T cell responses. It is possible that additional age-related differences in T cell responses will become more apparent over longer periods of time or that older individuals may experience deficits in the coordination of immune responses rather than differences in the relative frequencies of different cellular responses (Rydyznski Moderbacher et al. 2020). Our cohort was relatively young (median age of 43 years, with the upper quartile of 53 years), and it is possible that we lacked sufficient numbers of older patients to observe significant differences. Despite prior studies showing potential sex differences in early immune responses and immune activation during acute infection (Takahashi et al. 2020), sex appeared to have minimal impact on long-term SARS-CoV-2-specific T cell or antibody immune responses in our cohort, which was nearly 50% female.

There has been intense interest in understanding the cause and effects of persistent COVID-19 symptoms (Carfì et al. 2020; Hellmuth et al. 2021; Tenforde et al. 2020; Huang et al. 2021; Datta, Talwar, and Lee 2020). Similar to other reports (Sudre et al. 2020), we observed a higher proportion of women with persistent symptoms at the time of the first study visit (median 53 days after onset of symptoms), when nearly half of the entire cohort had one or more persistent symptoms attributed to COVID-19. As a result, we had a unique opportunity to study immune responses and inflammation in individuals with or without these symptoms and identified several unique patterns. Overall, we did not detect clear immunologic differences between individuals with and without persistent symptoms, even when they persisted 4 or more months following acute infection, suggesting that persistent symptoms and prolonged recovery is likely a multifactorial process not solely dependent on (or reflected by) adaptive immune responses measured in this study. Much larger studies that clearly define the diverse phenotypes of individuals with persistent symptoms and prolonged recovery will likely be needed to provide a mechanistic understanding of this important group.

We observed that antibody responses were strongly and positively associated with the frequency of S- and N-specific CD4+ T cells as measured by ICS. Most importantly, we observed the strongest correlations with antibody neutralizing capacity and virus-specific CD4+ responses across ICS and AIM assays. These results are consistent with other studies that have shown an association between the magnitude of SARS-CoV-2-specific CD4+ T cells and antibody levels (Grifoni et al. 2020; Rydyznski Moderbacher et al. 2020). These data suggest the potential importance of follicular helper T cell responses in lymph nodes, and further tissue-based analyses will be important to more clearly define the coordinated adaptive and humoral immune response in diverse populations.

Strengths of this study include the diversity of the cohort and the even distribution of individuals according to disease severity followed longitudinally over 8 months from initial symptoms. There are several notable limitations. First, the timeframe of sampling was limited to the recovery phase of COVID-19, and it is possible that there are clinically important biological correlates that could have been identified had samples from the infectious period been available. Further analyses leveraging this and other cohorts will assess these predictors. Second, while there is intense interest in understanding post-acute sequelae of SARS-CoV-2 infection, there remains no consensus case definition for this condition. It is possible that our case definition was highly sensitive but lacked the specificity needed to detect biological differences characterizing this condition. As clinical phenotypes of post-acute sequelae of COVID-19 become better-defined, more focused analyses could yield important results. Finally, this study involved a large number of analyses and comparator groups using assays with inherent intra- and inter-participant variation and a high degree of collinearity between study factors, making it difficult to make specific biological or causal inferences. As a result, we used a targeted approach to focus analyses on primary endpoints of interest, and we acknowledge our results are hypothesis generating and need to be confirmed in future studies and/or in other cohorts.

Nonetheless, we observed important unique patterns across the various assays measuring adaptive and humoral immune responses for various clinical factors such as initial clinical severity defined by hospitalization or ICU care and pre-existing pulmonary disease. These data suggest that apart from severity of initial illness, pre-existing medical conditions may have important influence over the longitudinal adaptive immune responses.

## METHODS

### LIINC Study

Participants were volunteers in the University of California, San Francisco-based Long-term Impact of Infection with Novel Coronavirus (LIINC) study. LIINC is an observational cohort that enrolls individuals with SARS-CoV-2 infection documented by clinical nucleic acid amplification testing who have recovered from the acute phase of infection. Volunteers are recruited by clinician- or self-referral. They are eligible to enroll between 14 and 90 days after onset of COVID-19 symptoms and are offered monthly visits until 4 months after illness onset; they are then seen every 4 months thereafter. At each study visit, participants undergo a detailed clinical interview that includes demographic information, COVID-19 diagnosis, illness, and treatment history, assessment of medical comorbidities and concomitant medications, and evaluation of ongoing symptoms and quality of life. At the first visit, participants were asked to assess their level of disability related to the worst point in their acute illness according to 3 measures on a 3-point scale: mobility, ability to perform self-care, and ability to perform routine work and household obligations. Biospecimens are collected and stored.

For the current study, we selected LIINC participants who had at least two time points available for analysis. We excluded participants with HIV infection. We randomly selected participants that experienced low initial disease severity (defined as 4 or fewer points on the symptom severity scale without hospitalization), moderate severity (5-7 points without hospitalization), and highly severe disease (greater than 7 points and/or hospitalized) in order to have a sample population representing a wide spectrum of initial disease severity. We also assessed whether participants had persistent symptoms that they attributed to COVID-19. We defined a persistent symptom as a symptom that was noted to be newly present during acute infection that remained present at follow-up.

### Ethics Statement

All participants sign a written informed consent and the study was approved by the University of California, San Francisco Institutional Review Board (IRB# 20-30479).

### PBMC Isolation

Whole blood was collected from EDTA tubes (COVID-19 participants) or from Buffy coats from healthy unexposed, anonymous donors collected and stored prior to November, 2019 in the San Francisco, Bay Area. Whole blood was centrifuged for 10 min at 1600 rpm to separate plasma. Plasma was then removed and stored at −80°C. Peripheral blood mononuclear cells (PBMCs) were isolated by density-gradient Ficol-Paque (GE Healthcare, Chicago, IL) using SepMate tubes (StemCell Technologies, Cambridge, MA). PBMC were cryopreserved in heat inactivated fetal bovine serum (Phoenix Scientific, Bangkok, Thailand) containing 10% DMSO (Sigma-Aldrich) and stored in liquid nitrogen.

### Peptide Pools

SARS-CoV-2 specific T cell peptide pools were purchased from Miltenyi Biotec (PepTivator SARS-CoV-2 Prot_S and PepTivator SARS-CoV-2 Prot_N) and resuspended in DMSO. SARS-CoV-2 S and N are pools of lyophilized peptides, consisting of 15 amino acids length with 11 amino acids overlap, covering the immunodominant sequence domains of the Spike (“S”) or Nucleocapsid (“N”) proteins of SARS-CoV-2.

### Activated Induced Cell Marker Assay

Cryopreserved PBMC were thawed in 10 ml of complete RPMI (Gibco) containing 10% fetal bovine serum and stimulated in complete RPMI containing 10% human AB serum (Sigma-Aldrich). Cells were then cultured for 24 hours in 96-wells U bottom plates at 1×10^6^ PBMC per well in the presence of either SARS-CoV-2 peptide pools (1 ug/ml), 10ug/ml phytohemagglutinin (PHA, Sigma-Aldrich) as positive control, or equimolar DMSO as negative control as previously described (Grifoni et al. 2020). All samples were analyzed on a BD LSR-II analyzer and analyzed with FlowJo X software. A complete list of antibodies are listed in **Supplementary Methods**. Whenever possible, longitudinal samples from individual participants were included in the same assay to minimize potential batch testing effects.

### Intracellular Cytokine Staining Assay

We implemented an in-house ICS assay as previously described with minor modifications to be consistent with other published SARS-CoV-2-specific assays (Henrich et al. 2017; Scully et al. 2018; Morley et al. 2019). Briefly, cells were rested overnight before stimulation. Cells were cultured for 8 hours at 37C in 96-wells U bottom plates at 1×10^6^ PBMC per well in presence of either SARS-CoV-2 peptides [1 µg/ml], CD3/CD28 beads (Gibco Dynabeads) as positive control, or equimolar DMSO as negative control. All conditions were in the presence of Golgi-plug containing brefeldin A (eBioscience), monensin (eBioscience), anti-CD28 (Biolegend, Clone CD28.2), and CD107a. After an 8-hour incubation, plates were put at 4°C overnight. The following day, Cells were washed and surfaced stained for 20 min at room temp in the dark. Following surface staining, cells were washed twice with PBS and then fixed/permeabilized (BD Cytofix/Cytoperm) for 45 min at 4°C in the dark. Cells were then washed twice with fix/perm wash buffer (BD Perm/Wash) and stained with intracellular antibodies for 45min at 4°C in the dark. A complete list of antibodies are listed in **Supplementary Methods**. All samples were analyzed on a BD LSR-II analyzer and analyzed with FlowJo X software. Whenever possible, longitudinal samples from individual participants were included in the same assay to minimize potential batch testing effects.

### Soluble Markers of Inflammation

Six different ELISA kits were used to detect the following markers, respectively: IL-6, IL-10, IP-10, D-Dimer, CD163, and CD14. Each kit was developed by a manufactured brand and standard protocol and dilutions were used in the preparation and analysis of each plate as follows: IL-6 ELISAs were prepared and conducted using the Millipore Sigma-Aldrich Human IL-10 ELISA Kit (Product Number RAB0306). The highest standard concentration used was 1000 pg/mL, decreasing concentration at a 3-fold dilution. Samples were run in duplicate without further dilution. IL-10 ELISAs were prepared and conducted using the Millipore Sigma-Aldrich Human IL-10 ELISA Kit. The highest standard concentration used was 150 pg/mL, decreasing concentration at a 2-fold dilution. Samples were run in duplicate without further dilution. D-Dimer ELISAs were conducted using the ThermoFisher Scientific Invitrogen Human D-Dimer ELISA Kit. The highest standard concentration used was 60 pg/mL, decreasing concentration at a 3-fold dilution. Samples were run in duplicate at a 250,000-500,000-fold dilution. IP-10 ELISAs were conducted using the Invitrogen Human IP-10 ELISA Kit. The highest standard concentration used was 500 pg/mL, decreasing concentration at a 2-fold dilution. Samples were run in duplicate without further dilution. CD163 ELISAs were prepared and conducted using the ThermoFisher Scientific Invitrogen Human CD163 (M130) ELISA Kit. The highest standard used was 8000 pg/mL, decreasing at a serial dilution of 40%. Samples were run in duplicate at a 50-fold dilution. CD14 ELISAs were prepared and conducted using the RnD Systems Quantikine ELISA Human CD14 Immunoassay. The highest concentration used was 8000 pg/mL, decreasing concentration at a 2-fold dilution. Samples were run in duplicate at a 200-250-fold dilution. Colorimetric changes were detected and quantified using the Spectramax M3 Plate Reader.

### SARS-CoV-2 Antibody Testing

Serum was tested for antibodies at UCSF using an in-house multiplex microsphere assay (Luminex platform) to detect IgG against SARS-CoV-2 spike, receptor binding domain (RBD), and two preparations of the N protein (on full length and one fragment). We used a published protocol with modifications (Wu et al. 2019). Plasma samples were diluted to 1:100 in blocking buffer A (1xPBS, 0.05% Tween, 0.5% bovine serum albumin (BSA), 0.02% sodium azide). Antigen concentrations used for bead coupling were as follows: S, 4 ug/mL; RBD, 2 ug/mL; and N, 3 ug/mL. Concentration values were calculated from the Luminex median fluorescent intensity (MFI) using a plate-specific standard curve from serial dilutions of a pool of positive control samples (https://github.com/EPPIcenter/flexfit). A cutoff for positivity was established for each antigen as the maximum concentration value observed across 114 pre-pandemic SARS-CoV-2 negative control samples tested on the platform.

### PhenoSense SARS CoV-2 nAb Assay

The measurement of nAb activity using the PhenoSense SARS CoV-2 nAb Assay (Monogram Biosciences, South San Francisco, CA) is performed by generating HIV-1 pseudovirions that express the SARS CoV-2 spike protein. The pseudovirus is prepared by co-transfecting HEK293 producer cells with an HIV-1 genomic vector that contains a firefly luciferase reporter gene together with a SARS CoV-2 spike protein expression vector. Neutralizing antibody activity is measured by assessing the inhibition of luciferase activity in HEK293 target cells expressing the ACE2 receptor and TMPRSS2 protease following pre-incubation of the pseudovirions with serial dilutions of the serum specimen.

Data are displayed by plotting the percent inhibition [% Inhibition = 100% – ((RLU(Pseudovirus+Sample+Cells)) ÷ (RLU(Pseudovirus+Diluent+Cells)) x 100%)] of luciferase activity expressed as relative light units (RLU) vs. the log_10_ reciprocal of the serum/plasma dilution. Neutralizing antibody titers are reported as the reciprocal of the serum dilution conferring 50% inhibition (ID50) of pseudovirus infection. To insure that the measured nAb activity is SARS CoV-2 nAb specific, each test specimen is also assessed using a non-specific pseudovirus (specificity control) that expresses a non-reactive envelope protein of an unrelated virus (e.g. avian influenza virus H10N7).

### Statistical Analyses

Flow cytometric, antibody and cytokine data were generated blinded to participant information. Comparisons of M0 values across comparator groups incorporated non-parametric Mann-Whitney or Kruskal-Wallis test with unadjusted Dunn correction for multiple comparisons using Prism v. 8 (GraphPad Software). Fisher’s exact test when any N<5 was used to compare tabular data. Spearman Rank Correlation analysis was used to compare T cell, antibody and soluble markers of inflammation (Prism). Whereas using all timepoints from all participants may lead to oversampling of some individuals, for correlations we used calculated weighted averages, representing the outcome of interest across all time points for each individual. Linear regression modeling was performed using SPSS v. 27 (IBM) including covariates of interest identified in the univariate analyses. For longitudinal analyses, linear mixed effects modelling was performed for each immunologic outcome (log transformed) in R (version 4.0) using lme4 package (version 1.1) with time and individual factors (e.g. Age, Sex, Ethnicity, Hospitalization, ICU admission, Symptoms at the first study time point and the month 4 time point, prior history of pulmonary disease) as predictors, and random effects based on participant. Sensitivity analyses were performed excluding month 8 data (when available) to rule out assay batch effects.

## Data Availability

The raw data supporting the conclusions of this article will be made available by the authors upon request. All data points are shown as individual values in the graphs.

## ACKNOWLEDGEMENTS

We are grateful to the LIINC study participants and to the clinical staff who provided care to these individuals during their acute illness period and recovery. We acknowledge LIINC study team members Tamara Abualhsan, Mireya Arreguin, Jennifer Bautista, Monika Deswal, Heather Hartig, Marian Kerbleski, Lynn Ngo, Fatima Ticas, and Meghann Williams for their contributions to the program. We are grateful to Khamal Anglin, Grace Bronstone, Jessica Chen, Michelle Davidson, Kevin Donohue, Peyton Ellis, Sarah Goldberg, Scott Lu, Jonathan Massachi, Sujata Mathur, Irum Mehdi, Victoria Wong Murray, Enrique Martinez Ortiz, Jesus Pineda-Ramirez, Mariela Romero, Paulina Rugart, Hannah Sans, Joshua Shak, Jaqueline Tavs, and Jacob Weiss for assistance with data entry and validation. We thank Shane Crotty and Alessandro Sette for providing experimental details and advice regarding the activation marker induced assay. We thank the UCSF Core Immunology Laboratory for processing some samples. We are grateful to the UCSF AIDS Specimen Bank for specimen processing and for managing the LIINC biospecimen repository.

## Contributions

MJP, AND, FTA, RLR, TJH designed the study, which was supported through funding to MS, MG, JDK, JNM, SGD, and TJH. MJP, LT, RH, VT, YH, and EF collected clinical data and biospecimens. LT, CCN, NSI, SEM, JD, CT, JH, KT, and OJ performed PBMC and plasma isolation and storage. Specimens were analyzed by LT, CCN, NSI, SEM, JD, JH, KT, and OJ in the laboratories of BG and TJH. LT, TW, and CJP performed and interpreted the neutralization assays. MJP, AND, ST, FTA, RLR, and TJH performed and/or interpreted the statistical analyses. MJP, AND, RLR, and TJH drafted the initial manuscript. All authors edited, reviewed, and approved the final manuscript.

