## Supplementary Materials for "Long-Term SARS-CoV-2-Specific Immune and Inflammatory Responses Across a Clinically Diverse Cohort of Individuals Recovering from COVID-19"

**AIM DMSO Control**

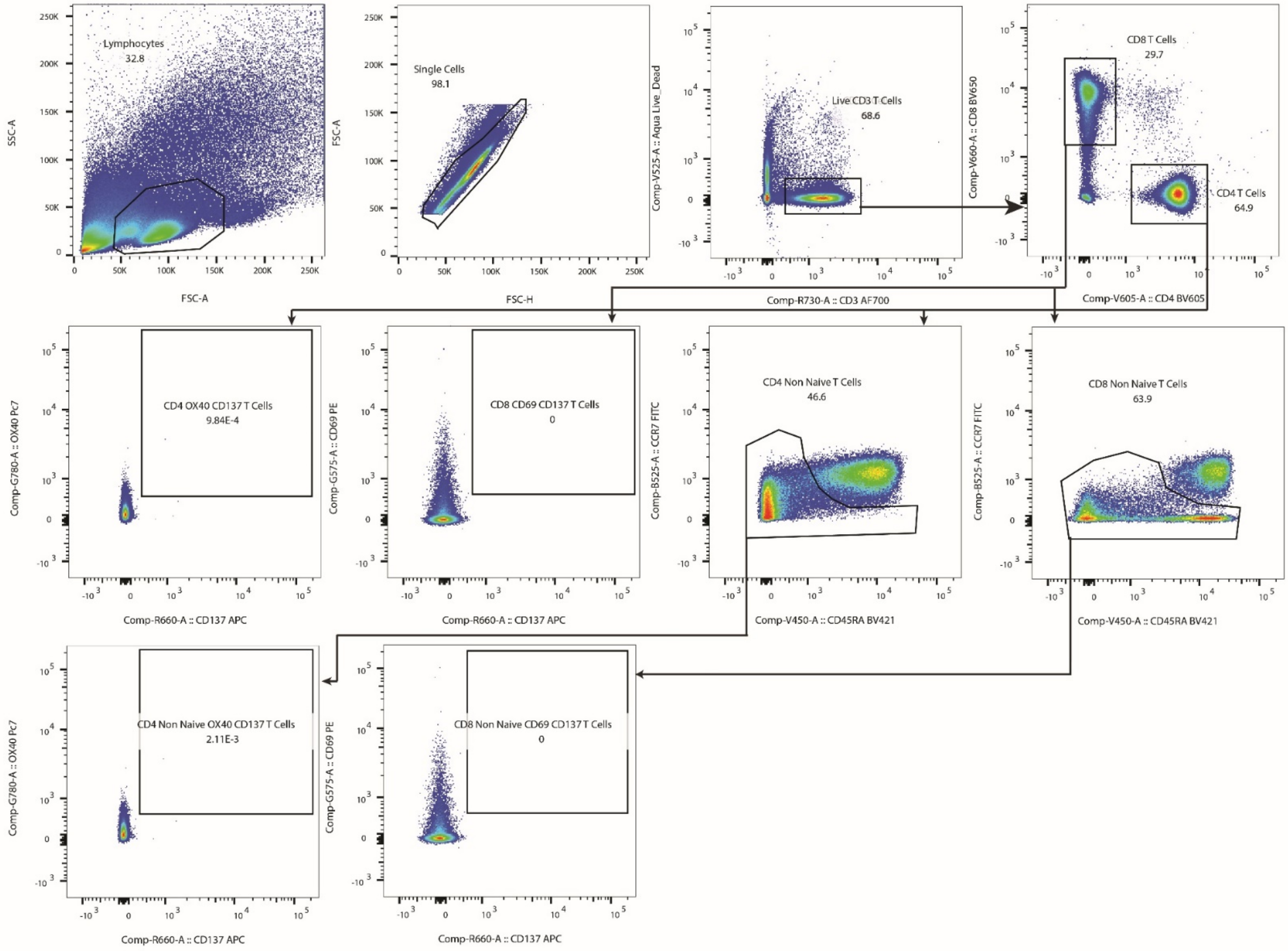

z

**Supplementary Figure 1.** Gating strategy for the Activation Induced Marker (AIM) assay. Data from memory (non naïve) CD4 and CD8 T cells were used in the primary analyses.

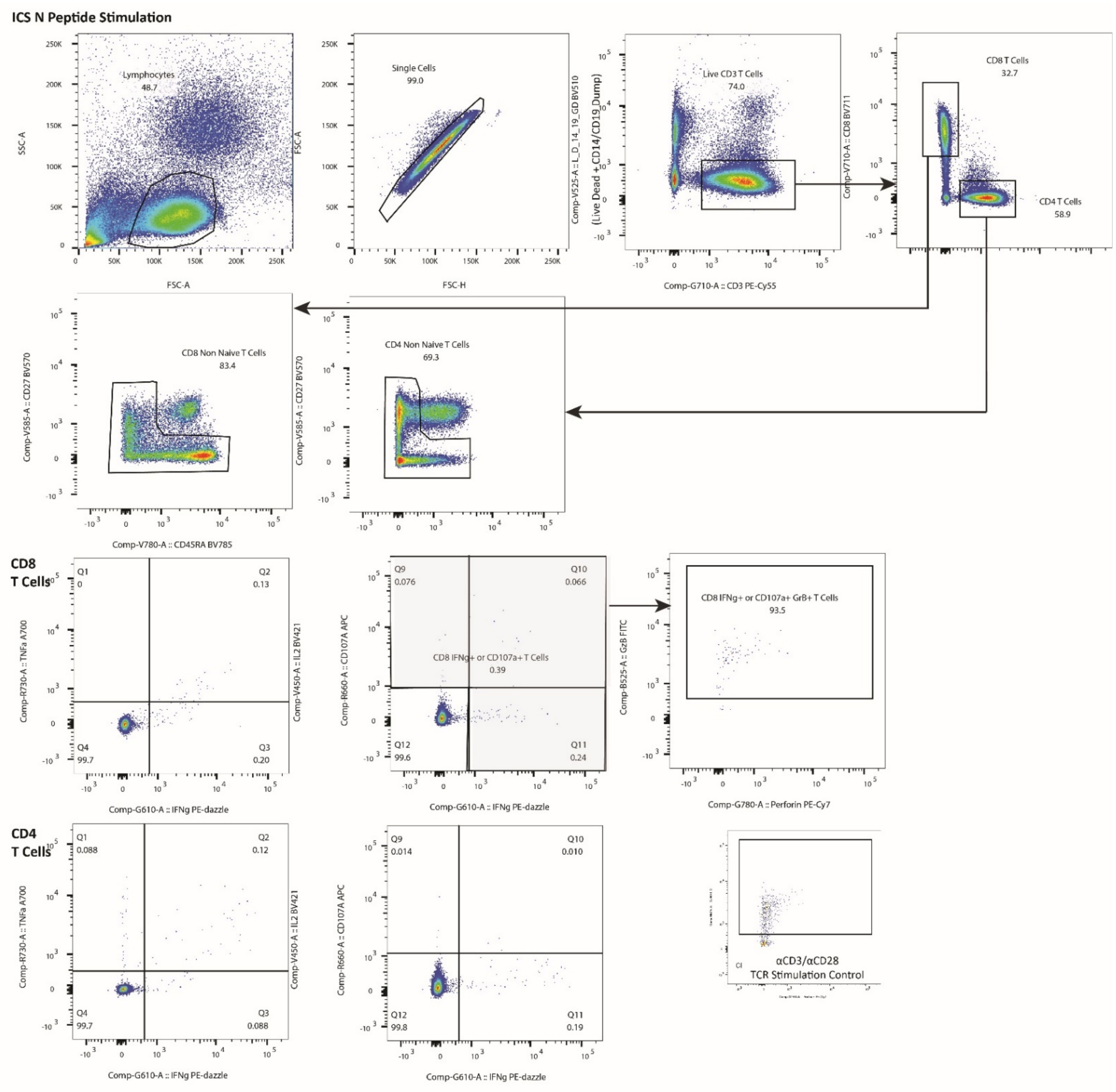

**Supplementary Figure 2.** Gating strategy for the Intracellular Cytokine Staining (ICS) assay. Data from memory (non naïve) CD4 and CD8 T cells were used in the analyses.

**Supplementary Table 1.** Participants with Activation Induced Marker (AIM) and Intracellular Cytokine Staining (ICS) Assay Results Greater than Pre-COVID-19 Control Samples Across All Timepoints.

| Assay | N[a] | N Above Upper Interquartile Range (%+ Cells) of Pre-COVID-19 Controls (% of N) |
|---|---|---|
| CD4 AIM or ICS (N or S) | 69 | 69 (100) |
| CD8 AIM or ICS (N or S) | 69 | 66 (95.7) |
| CD4 AIM N or S | 68 | 67 (98.5) |
| CD4 AIM N | 68 | 67 (98.5) |
| CD4 AIM S | 68 | 65 (95.6) |
| CD8 AIM N or S | 68 | 52 (76.5) |
| CD8 AIM N | 68 | 46 (67.6) |
| CD8 AIM 2 | 68 | 41 (60.3) |
| CD4 ICS N or S (IFNγ+/TNFα+ or IFNγ+) | 65 | 65 (100) |
| CD4 ICS N | 64 | 64 (100) |
| CD4 ICS S | 64 | 63 (98.4) |
| CD8 ICS N or S (IFNγ+/TNFα+ or IFNγ+) | 65 | 53 (81.5) |
| CD8 ICSN | 65 | 47 (72.3) |
| CD8 ICS S | 63 | 46 (73.0) |

[a] Number of participants from the total analysis cohort (N=70) with valid AIM or ICS assay results (*e.g*. sufficient numbers of cells for analysis, adequate staining)

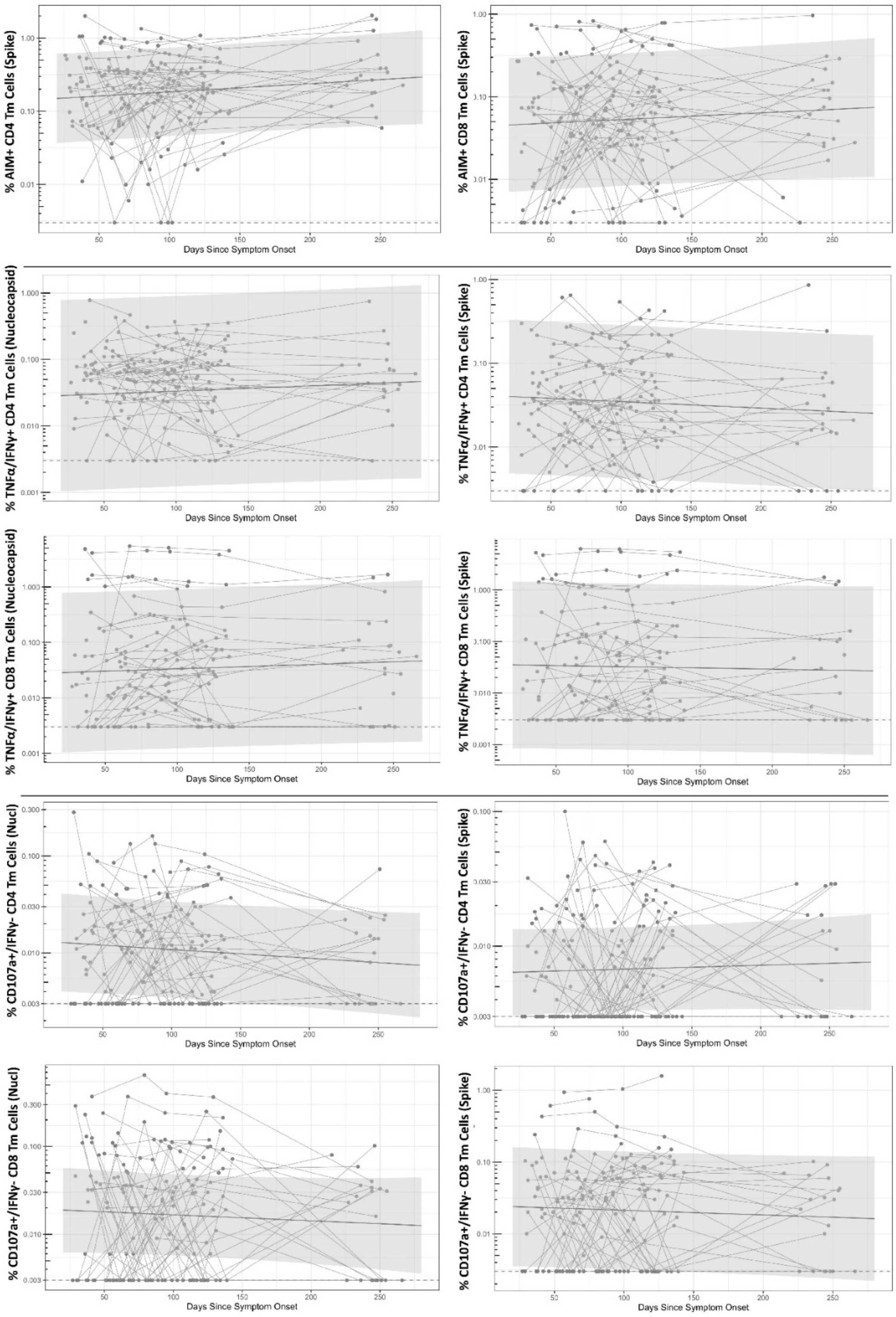

% AIM+ CD4 Tm Cells (Spike)
Days Since Symptom Onset
% AIM+ CD8 Tm Cells (Spike)
Days Since Symptom Onset
% TNFα/IFNγ+ CD4 Tm Cells (Nucleocapsid)
Days Since Symptom Onset
% TNFα/IFNγ+ CD4 Tm Cells (Spike)
Days Since Symptom Onset
% TNFα/IFNγ+ CD8 Tm Cells (Nucleocapsid)
Days Since Symptom Onset
% TNFα/IFNγ+ CD8 Tm Cells (Spike)
Days Since Symptom Onset
% CD107a+/IFNγ- CD4 Tm Cells (Nucl)
Days Since Symptom Onset
% CD107a+/IFNγ- CD4 Tm Cells (Spike)
Days Since Symptom Onset
% CD107a+/IFNγ- CD8 Tm Cells (Nucl)
Days Since Symptom Onset
% CD107a+/IFNγ- CD8 Tm Cells (Spike)
Days Since Symptom Onset

**Supplementary Figure 3.** Longitudinal T cell responses as measured by AIM SARS-CoV-2 Spike assay (**A**), dual expression of IFNγ+ and TNFα CD8+ and CD4+ T cells by ICS (**B**), and expression of CD107a (IFNγ negative) CD4+ and CD8+ T cells (**C**). Solid line and shaded region represent the median model prediction and 95% prediction interval from linear mixed effects modeling. Dashed lines represent assay limits of detection.

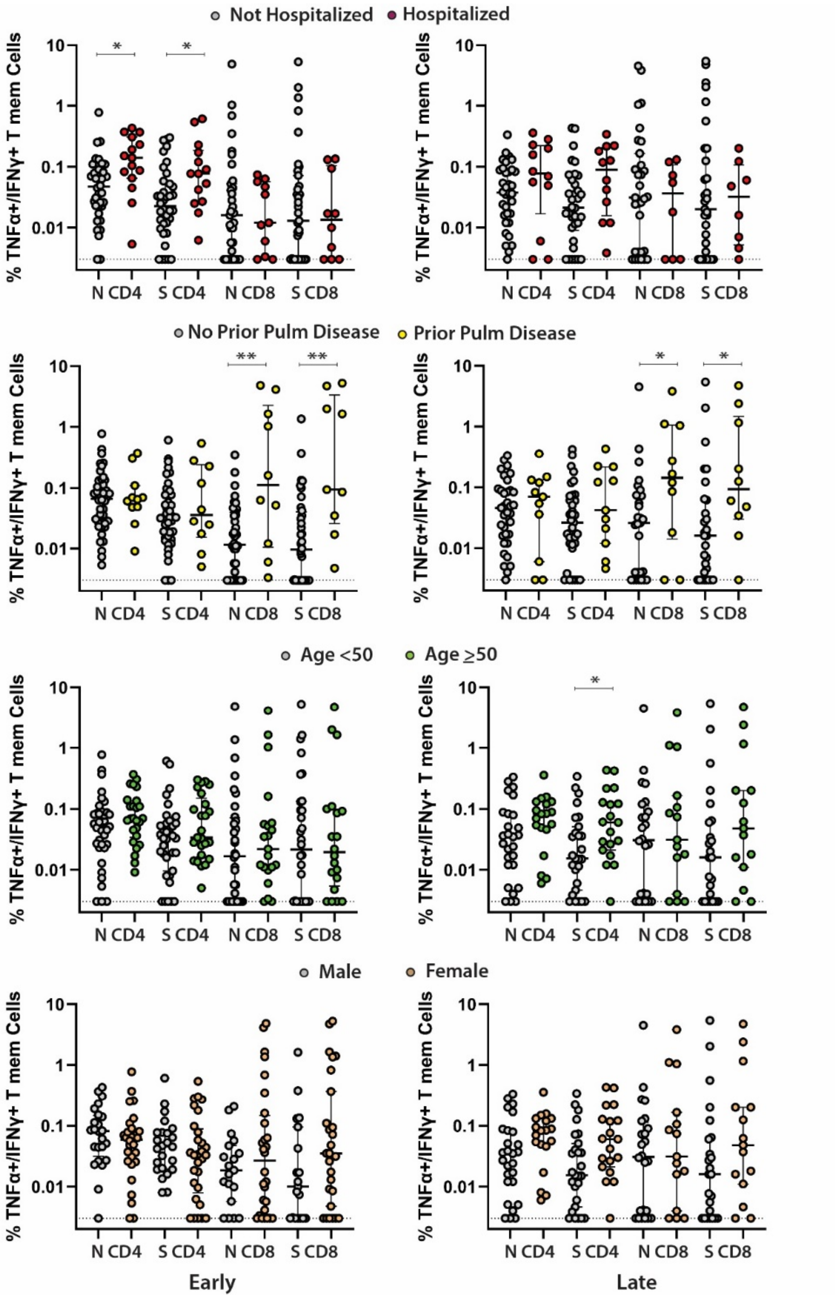

**Supplementary Figure 4.** Frequency of SARS-CoV-2-specific TNFα+/IFNγ+ T cell CD4+ T cells as measured by the ICS assay in study participants for various clinical and demographic factors at early (left panels, median 53 days after onset of symptoms) versus late (right panels, median 123 days from onset of symptoms) cross-sectional analysis time points. All data points are shown as individual points. Bars and lines in cross sectional data represent median values and interquartile ranges; (*) P<0.05, (**) P<0.01, (***) P<0.001 by non-parametric analyses.

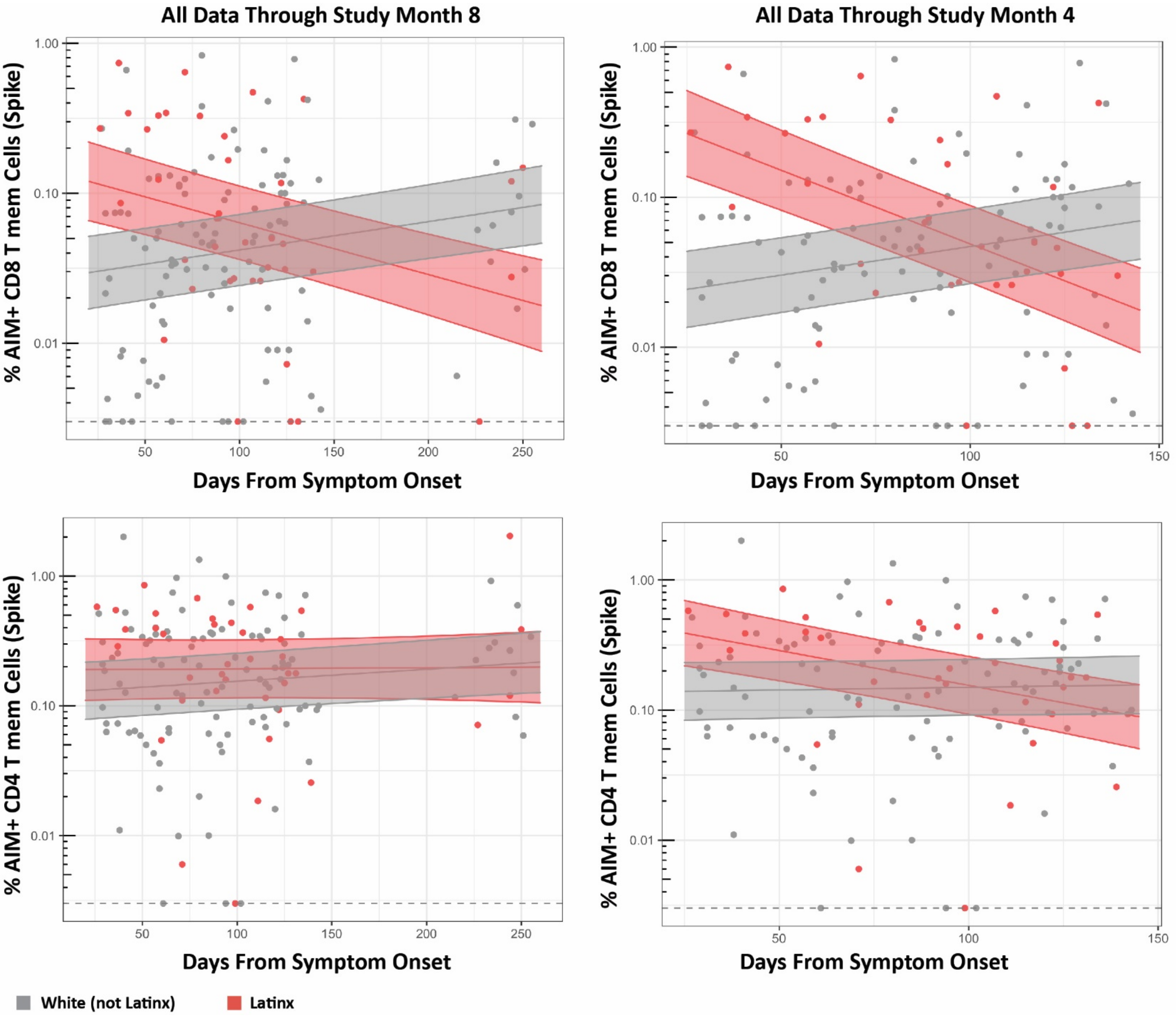

**Supplementary Figure 5.** Mixed effects models of longitudinal AIM+ CD8 T cell (top) and AIM + CD4 T cell (bottom) responses grouped by Latinx and white (non Latinx) ethnicity for all data through the month 8 collection time points are shown. In order to rule out potential batch effects, mixed effects models were similar including only data from month 4 time points (right panels). Solid line and shaded region represent the median model prediction and 50% prediction interval from linear mixed effects modeling. Dashed lines represent assay limits of detection.

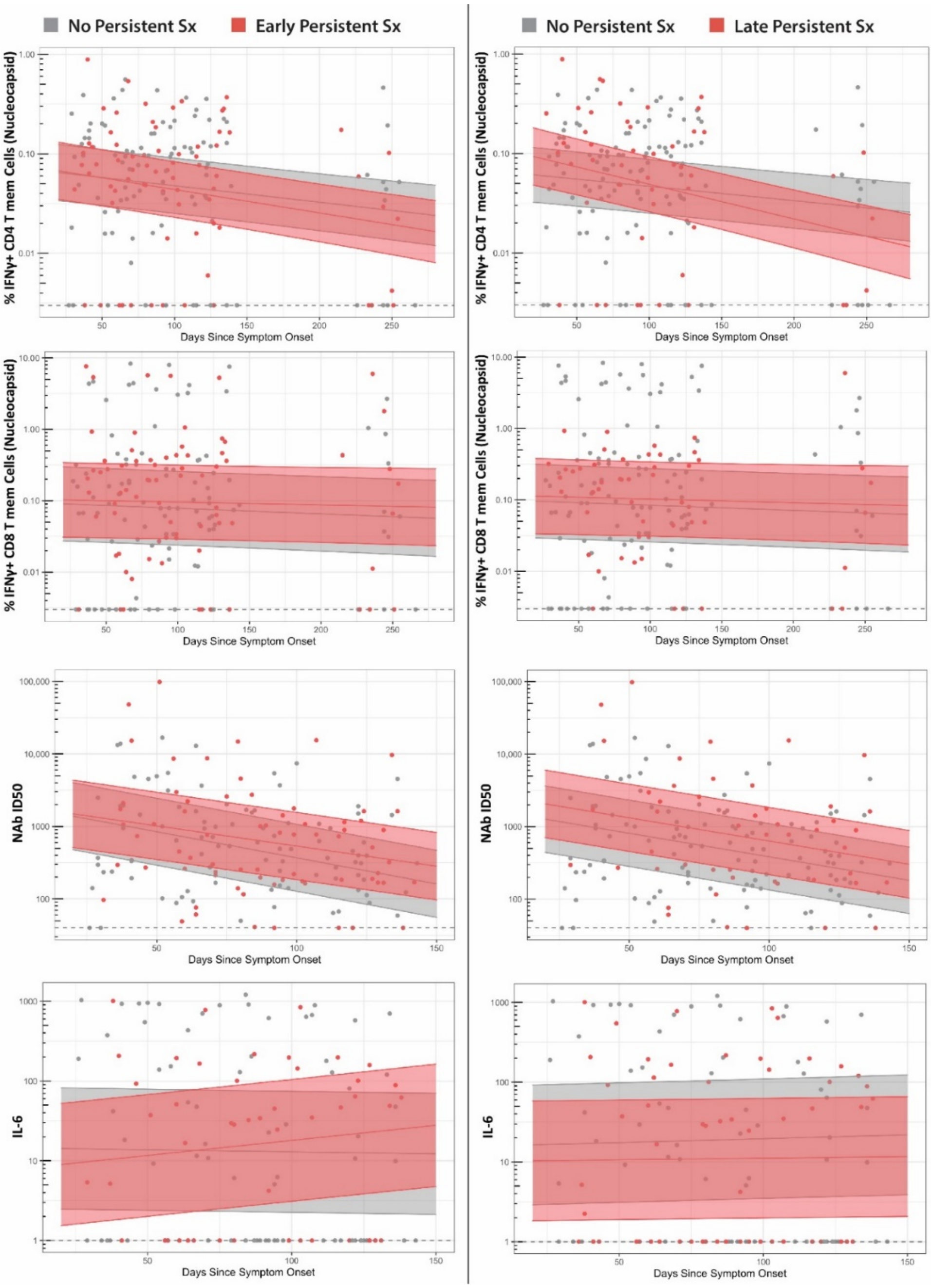

**Supplementary Figure 6**. Minimal differences in T cell, neutralizing antibody and inflammatory markers were observed between those with persistent following acute infection at the first study time point (left panels, median 53 days) and approximately 4 months after onset of acute illness (right panels, median 123 days). Solid line and shaded region represent the median model prediction and 50% prediction interval from linear mixed effects modeling. Dashed lines represent assay limits of detection. NAb = neutralizing antibody.

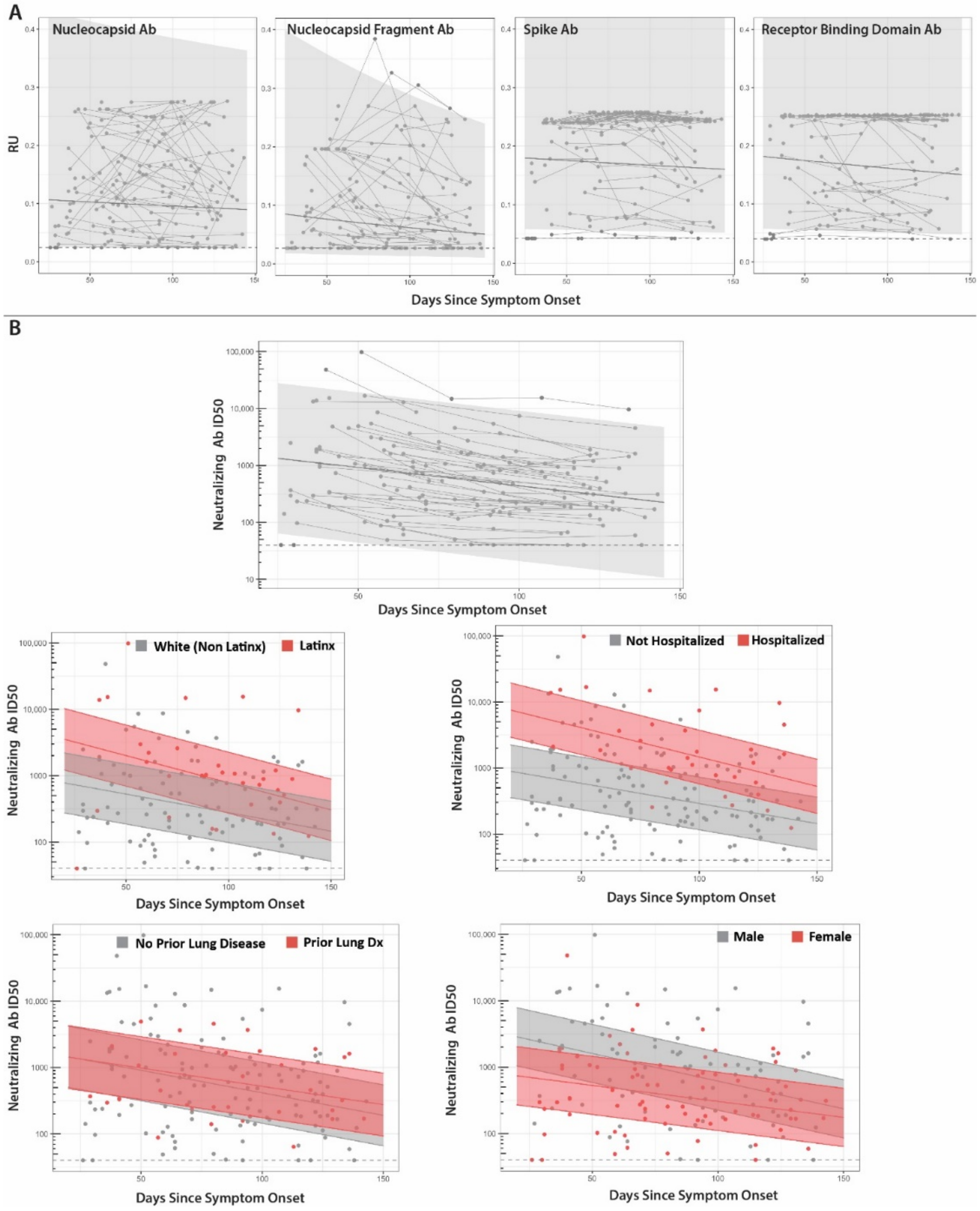

**Supplementary Figure 7**. Longitudinal antibody response (relative light units; RU) data are shown for data generating using the Luminex IgG assay across the nucleocapsid (N), nucleocapsid fragment (N.361), spike (S) and receptor binding domain (RBD) are shown in the top panels (**A**). Neutralizing capacity (**I**nfectious Dose, 50% [ID50] of Spike pseudovirus in presence of participant serum) for all participants and by various clinical and demographic factors are shown in the bottom panels (**B**). Points and connecting lines represent raw data for each individual. Solid line and shaded region represent the median model prediction and 95% prediction interval from linear mixed effects modeling (50% prediction intervals for comparisons between factors). Dashed lines represent assay limits of detection.

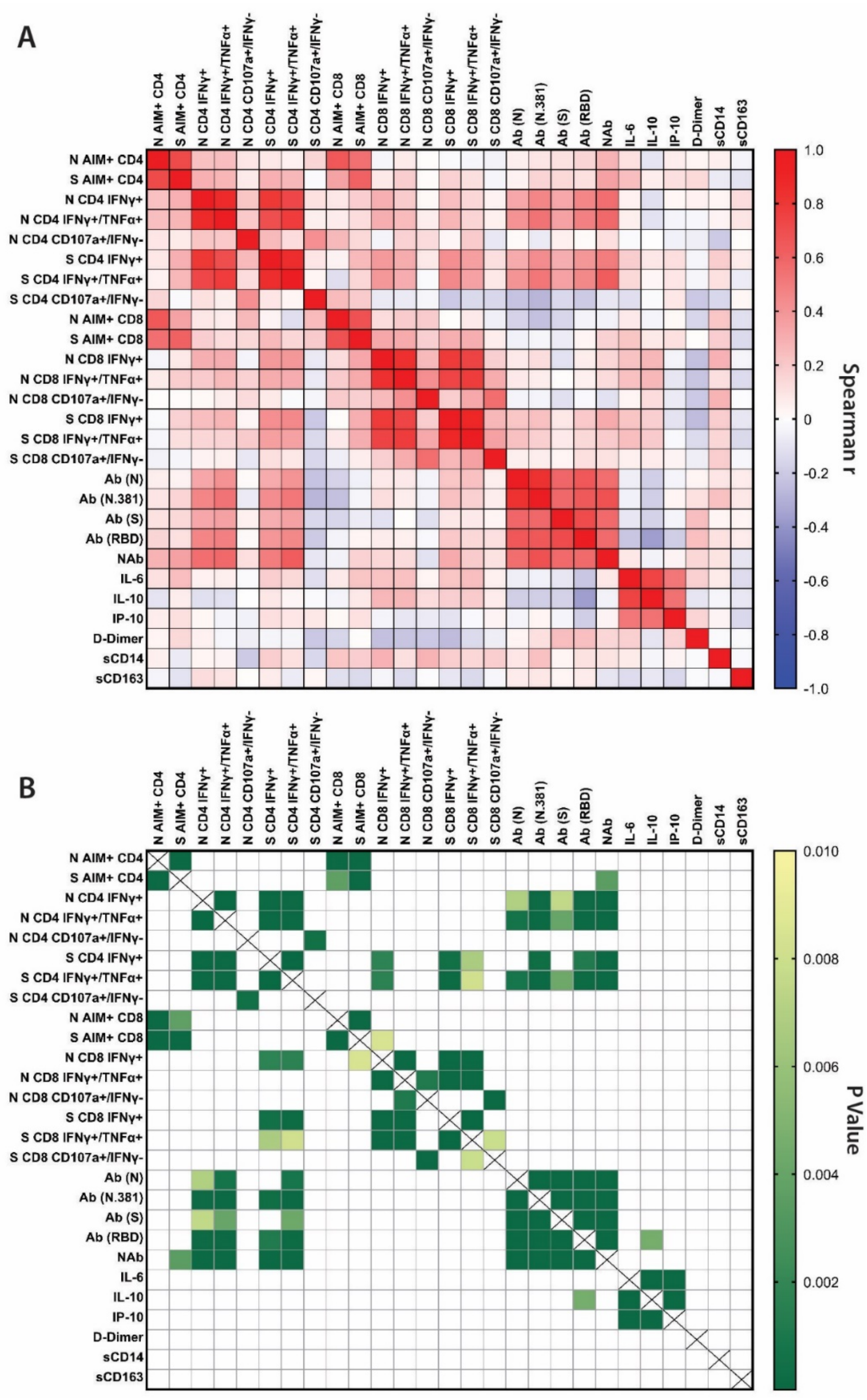

**Supplementary Figure 8**. Spearman correlation matrix of all T cell, antibody and inflammatory marker data is shown in (**A**). P value heatmap is shown in (**B**).

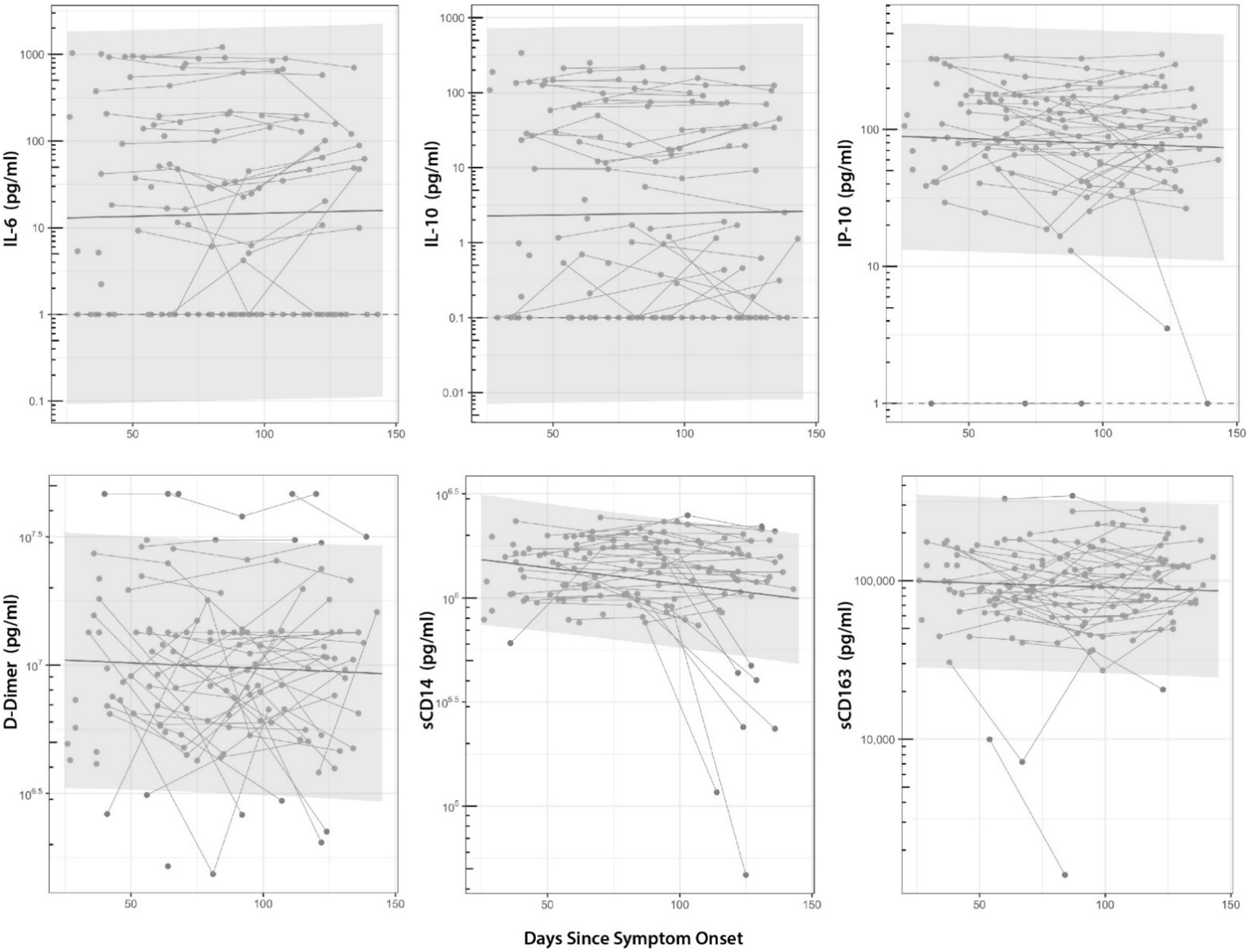

**Supplementary Figure 9.** Circulating markers of cytokines and markers of inflammation over time. With the exception of a modest decrease in sCD14 over time (1pg/ml per day (P=0.006), levels remained constant over 4 months following onset of COVID-19 symptoms. Solid line and shaded region represent the median model prediction and 95% prediction interval from linear mixed effects modeling

**Supplementary Table 2.** Antibodies Included in the Activation Induced Marker (AIM) Assay

| Antibody | | Fluorochrome | Clone/vendor/catalog |
|---|---|---|---|
| 1 | CD45RA | BV421 | HI100/Biolegend/304130 |
| 2 | CD14 | V500 | M5E2/BD/561391 |
| 3 | CD19 | V500 | HIB19/BD/561121 |
| 4 | Live/Dead | Aqua | Invitrogen/L34966 |
| 5 | CD8 | BV650 | RPA-T8/BioLegend/301042 |
| 6 | CD4 | BV605 | RPA-T4/BD/562659 |
| 8 | CCR7 | FITC | G043H7/Biolegend/353216 |
| 9 | CD69 | PE | FN50/BD/555531 |
| 10 | OX40 | PE-Cy7 | Ber-ACT35/Biolegend/350012 |
| 11 | CD137 | APC | 4B4-1/BioLegend/309810 |
| 12 | CD3 | AF700 | OKT3/Biolegend/317340 |
| 13 | CD40L | PE-Dazzle | 24-31/Biolegend/310840 |

**Supplementary Table 3.** Antibodies Included in the Intracellular Cytokine Staining (ICS) Assay

| Antibody | | Fluorochrome | Clone/vendor/catalog |
|---|---|---|---|
| 1 | IL-2 | BV421 | MQ1-17H12/Biolegend/500328 |
| 2 | CD14 | BV510 | M5E2/Biolegend/301842 |
| 3 | CD19 | BV510 | HIB19/Biolegend/302242 |
| 4 | TCR g/d | BV510 | B1/Biolegend/331220 |
| 5 | Live/Dead | Aqua | Invitrogen/L34966 |
| 6 | CD27 | BV570 | O323/BioLegend/302825 |
| 7 | CD4 | BV650 | OKT4/Biolegend/317436 |
| 8 | CD8 | BV711 | SK1/Biolegend/344733 |
| 9 | CD45RA | BV785 | HI100/Biolegend/304140 |
| 10 | GranB | FITC | Ber-ACT35/Biolegend/350012 |
| 11 | IFNg | PE-CF594 | GB11/Biolegend/515403 |
| 12 | CD3 | PE-Cy5.5 | SK7/eBioscienc/35-0036-42 |
| 13 | CD107a | APC | H4A3/Biolegend/328620 |
| 14 | TNFa | AF700 | 6402/R&Dsystems/IC9677N-100UG |
